# Chronicling menstrual cycle patterns across the reproductive lifespan with real world data

**DOI:** 10.1101/2024.01.09.24301041

**Authors:** Adam C. Cunningham, Lubna Pal, Aidan P. Wickham, Carley Prentice, Frederick G. B. Goddard, Anna Klepchukova, Liudmila Zhaunova

**Author notes:** **Correspondence should be addressed to:** Adam Cunningham, 27 Old Gloucester Street, London, WC1N 3AX, United Kingdom.

## Abstract

**Background:** The intricate hormonal and physiological changes of the menstrual cycle can influence health on a daily basis. Although prior studies have helped improve our understanding of the menstrual cycle, they often lack diversity in populations included, sample size, and the span of reproductive and life stages. This paper aims to describe the dynamic differences in menstrual cycle characteristics and associated symptoms by age in a large global cohort of period tracking application users. This work aims to contribute to our knowledge and understanding of female physiology at varying stages of reproductive aging.

**Methods:** This cohort study included self-reported menstrual cycle and symptom information in a sample of Flo application users aged 18-55. Cycle and period length and their variability, and frequency of menstrual cycle symptom logs are described by age of user.

**Results:** Based on data logged by over 19 million global users of the Flo app, the length of the menstrual cycle and period show clear age associated patterns. With higher age, cycles tend to get shorter (Cycle length: D̄ = 1.85 days, Cohen’s D= 0.59) and more variable (Cycle length SD: D̄ = 0.42 days, Cohen’s D= 0.09), until close to the chronological age (40-44) suggesting menopausal transition, when both cycles and periods become longer (Cycle length: D̄ = 0.86 days, t=48.85, Cohen’s D=0.26; Period length: D̄ =0.08, t=15.6, Cohen’s D=0.07) and more variable (Cycle length SD: D̄ =2.80 days, t=111.43, d=0.51; Period length SD: D̄ =0.23 days, t=67.81, Cohen’s D=0.31). The proportion of individuals with irregular cycles was highest in participants aged 51-55 (44.7%), and lowest in the 36-40 age group (28.3%). The spectrum of common menstrual cycle related symptoms also varies with age. Frequency of logging of cramps and acne is lower in older participants, while logs of headache, backache, stress and insomnia are higher in older users. Other symptoms show different patterns, such as breast tenderness and fatigue peaking between the ages of 20-40, or mood swings being most frequently logged in the youngest and oldest users.

**Conclusion:** The menstrual cycle, and related symptoms are not static throughout the lifespan. Understanding these age related differences in cycle characteristics and symptoms is important in understanding how best to care for and improve daily experience for menstruators across the reproductive life span.

## Introduction

Menstruation, and menstrual health concerns are severely underrepresented in the health research landscape^1^. The female reproductive system ages faster than other systems in the body^2^. This has profound implications for the menstrual cycle, fertility, and the rest of the body and leads to female reproductive lifespan being shorter than the average expected lifespan of the individual. The characteristics of the menstrual cycle, like cycle length and regularity, and the profile or severity of symptoms such as weight gain, sleep problems or headaches are highly age dependent^3^.

Aside from cycle and period length variability due to health conditions or other environmental exposures, it is known that cycle length (the interval in days between the onset of a menstrual bleed and the onset of the subsequent menstrual bleed) and period length (days of bleeding) tend to shorten and become more variable with age until the final menstrual period at menopause, when the menstrual cycle ceases completely^4^. However, this linear overview of menstruation and age may be too simplistic, and can disguise more subtle age related effects, particularly early and late in the reproductive life span, such as an increase in cycle length after age 50, in the years immediately before the menopause^5^. While large scale studies exist, they have often relied on participants recruited from specific settings, such as healthcare, or on relatively narrow age ranges. There is therefore a lack of large-scale research that investigates the cycle and related symptoms across the whole reproductive life span, in naturalistic samples.

Alongside differences in the length and variability of the menstrual cycle with advancing age, there are also differences in the menstrual cycle related symptoms experienced by individual menstruators. For example, breast tenderness is most common in the teenage years, but most severe in the late 30s and early 40s^6^. Reports of sleep disturbances tend to increase with advancing age, particularly during the phase of menopausal transition and early menopause^7^. Indeed, the transition to menopause is associated with a range of symptoms including hot flashes, mood changes, impaired cognition, sleep disturbance among others ^8^. Around 60-80% of women transitioning towards and into menopause experience classic menopause symptoms that are severe enough to lead to many seeking medical care^8–10^. Regardless of life stage, the average human female experiences approximately 400 cycles over their lifetime^11^, meaning that any disruptive menstruation related symptoms can lead to a significant health and quality of life burden.

Despite efforts to improve menstrual health literacy, understanding of what constitutes as a “normal” menstrual cycle, both from perspectives of menstruators and healthcare professionals remains wanting ^11^. For example, irregular length cycles may be expected in individuals beyond age 45 and regarded as a symptom of the transition to menopause, but would be a reason for medical investigation in younger aged individuals. Furthermore, many women are lacking clarity on when they should seek medical help for menstrual cycle related symptoms such as dysmenorrhea, which affects from 45-95% of menstruating individuals, depending on the classification criteria used ^12^, or for heavy menstrual bleeding which has a prevalence of around 27-39%^13^. Strikingly, it has been suggested that as few as 1 in 10 women in the UK felt they had received enough information about what to expect during the transition to menopause^14^. Improving our knowledge and understanding of the variability in symptoms experienced across the reproductive lifespan is necessary to help improve menstrual health literacy.

There are now many smartphone applications (mobile health *apps* [mHealth *app]*) available, and are increasingly being utilized towards tracking of the menstrual cycle in order to gain clarity on menstrual patterns and symptoms. These *apps* allow users to input data well beyond the actual menstrual events, including data on physical and mood symptoms, sexual and physical activity. Tracking the menstrual cycle can help users understand their physiology, and better plan for their biological goals as well as promote healthy habit forming or behavior ^15–17^.

Flo (by Flo Health UK Limited) is a health, well-being, and period tracker mHealth app for women and people who menstruate, with over 58 million monthly active users. The Flo app (https://flo.health/) offers artificial intelligence (AI) based period prediction allowing users the ability to track their periods, fertile window, and timing of ovulation as well as record a wide variety of physical and psychological symptoms. A unique aspect of The Flo *app* is that it allows users access to a large library of evidence-based educational content relating to menstrual and reproductive health, information that has been vetted by medical and scientific experts in the field of women’s health. The real-life data that can be collected through *apps* such as Flo offers the potential to serve as a rich resource for population health studies into menstrual cycle and its symptoms, reproductive wellness and disorders using ‘digital epidemiology’ approaches^18–20^.

In this paper we aim to describe differences in menstrual cycle length, bleeding length and symptom logged by a sample of women and people who menstruate aged 18-55 who have tracked their menstrual cycle and symptoms using the Flo *app*.

## Methods

This study included global users of the Flo *app* who recorded bleeding days or symptoms in a 12 month period between the 11th of December 2022 and the 11th of December 2023. Users were between 18-55 years of age at the time of data extraction, had not recorded a pregnancy during this time, and had recorded at least three menstrual cycles. Menstrual cycles that were shorter than 10 days, were excluded as unrealistic cycle lengths. To attempt to target users not on hormonal contraception, users who had set contraception reminders were also excluded. Cycle and period lengths were calculated based on self-reported user data. Users report the start and end dates of their period, with the cycle start date defined as the first day of bleeding. The period was defined as the number of days from the first day of bleeding and the last day of bleeding. If a user logs the start of bleeding, but does not provide an end date, the bleeding length is set to 5 days automatically. Participants provided informed electronic consent for aggregated data that could not identify them as individuals to be used for research purposes when agreeing to the usage terms and Privacy Policy of the Flo app. The study protocol was approved by an independent ethics review board (Western Copernicus Group Independent Review Board - WCG IRB, number 20226050).

Symptoms of interest were self-reported and included somatic (Cramps, Tender Breasts, Headache, Fatigue, Backache, Acne), gastrointestinal (Bloating, Constipation, Diarrhea, Nausea), mood (negative [Mood swings, Anxious mood, Depressed mood, Confused mood, Insomnia, Stress, Obsessive Thoughts], positive [Energetic mood, Happy]) and vaginal discharge (Sticky, Egg white, Creamy, Watery). Flo *app u*sers can record as many symptoms in each category as desired each day.

In addition to the cycle and symptom information and current age (in years) at time of data extraction were also extracted.

### Statistical Methods

Individual mean and standard deviations of cycle length and period length were calculated for each user. Standard deviation of cycle length was used to assess cycle variability. No outlier removal other than excluding cycles shorter than 10 days was performed as we expect longer and more irregular cycles in older age groups. These mean and standard deviation values were then aggregated by age group (18-25, 26-30, 31-35, 36-40, 41-45, 46-50 or 51-55), to facilitate comparisons between age groups. T-tests were used to compare differences between mean lengths and standard deviations between specific age groups. In addition, the mean cycle or period length, and mean standard deviation of cycle length was plotted by age to visualize differences in these outcomes by age.

Flo *app* users were classed as having irregular cycles if the difference between the longest and shortest cycle recorded within the time period of interest was greater than 7 days at least twice in the 12-month study period.

To investigate frequency of symptom logging by age, the mean number of symptom logs for each symptom type was plotted by age. For the symptom analysis, users who recorded more than five times the standard deviation of the mean of the total logs per user were excluded from the symptom level analysis due to having an extremely high rate of logging.

Due to the very large sample sizes used in these analyses, we do not report p-values. In large samples, p-values can be very small even if the difference between groups are very small, and not clinically useful or significant^21^. Instead we report mean differences and effect sizes where relevant.

All analysis was performed in R version 4.3.0 ^22^ and the following R packages: arrow v. 12.0.1^23^, arsenal v. 3.6.3^24^, gtsummary v. 1.7.1 ^25^ mgcv v. 1.8.42^26^, tidymv v. 3.4.2^27^, pwr v. 1.3.0^28^, patchwork v. 1.1.2 ^29^, tidyverse v. 2.0.0 ^30^ and rstatix v. 0.7.2^31^.

## Results

This study included 19,266,573 users aged 18-55 who had recorded at least three cycles longer than 10 days, had not used the pregnancy mode in the data extraction period and had not set reminders for contraception. Most users were in the 18-25 age group (N = 8,757,345), with a lower representation of users for each age group that follows (Table 1). Most users had a BMI of between 18.5-24.9,

**Table 1.** Summary statistics for the users included in the study.

|  | **Overall, N =**<br>**19,266,573** | **18-25, N =**<br>**8,757,345** | **26-30, N =**<br>**3,822,121** | **31-35, N =**<br>**2,948,002** | **36-40, N =**<br>**2,055,795** | **41-45, N =**<br>**1,234,400** | **46-50, N =**<br>**412,761** | **51-55, N**<br>**= 36,149** |
| --- | --- | --- | --- | --- | --- | --- | --- | --- |
| **Cycle Length in days, mean (SD)** |  |  |  |  |  |  |  |  |
| Mean Cycle Length | 28.54 (3.24) | 28.99 (3.31) | 28.76 (3.23) | 28.28 (3.11) | 27.68 (2.96) | 27.18 (2.81) | 27.15 (2.94) | 28.00 (3.70) |
| Standard Deviation of Cycle Length | 4.01 (4.74) | 4.14 (4.73) | 3.95 (4.81) | 3.80 (4.75) | 3.72 (4.65) | 3.92 (4.57) | 4.72 (4.89) | 6.52 (6.18) |
| **Period Length, mean (SD)** |  |  |  |  |  |  |  |  |
| Mean Period Length | 5.20 (0.92) | 5.28 (0.86) | 5.20 (0.94) | 5.13 (0.96) | 5.08 (0.98) | 5.06 (1.01) | 5.07 (1.04) | 5.13 (1.10) |
| Standard Deviation of Period Length | 0.65 (0.64) | 0.64 (0.64) | 0.65 (0.63) | 0.65 (0.63) | 0.66 (0.64) | 0.70 (0.67) | 0.76 (0.74) | 0.87 (0.84) |
| **Irregular Cycles, n (Row %)** |  |  |  |  |  |  |  |  |
| Irregular | 3,438,506 (18%) | 1,770,289 (20%) | 616,498 (16%) | 415,943 (14%) | 283,846 (14%) | 217,357 (18%) | 118,163 (29%) | 16,410 (45%) |
| Regular | 15,828,067 (82%) | 6,987,056 (80%) | 3,205,623 (84%) | 2,532,059 (86%) | 1,771,949 (86%) | 1,017,043 (82%) | 294,598 (71%) | 19,739 (55%) |

### Cycle Length

We found that mean cycle length shortens from 28.54 days in the 18-25 year old age band, to 27.15 days in the 46-50 year old age band? (Mean difference = 1.85 days, t=363.19, Cohen’s d=0.59) (Table 1). However, cycle length tends to increase after age 45, with the mean cycle length being longer in the 51-55 age group compared to the 46-50 age group (Mean difference = 0.86 days, t=48.85, Cohen’s d=0.26). A similar, non-linear pattern of cycle length differences is observed when plotting mean cycle length by year of age (Figure 1A). Mean cycle length increases from age 18 to around 21-22, when cycle length is longest. Between the ages of 22 and 45, cycle length decreases to its shortest, before increasing again post age 45.

**Figure 1.**
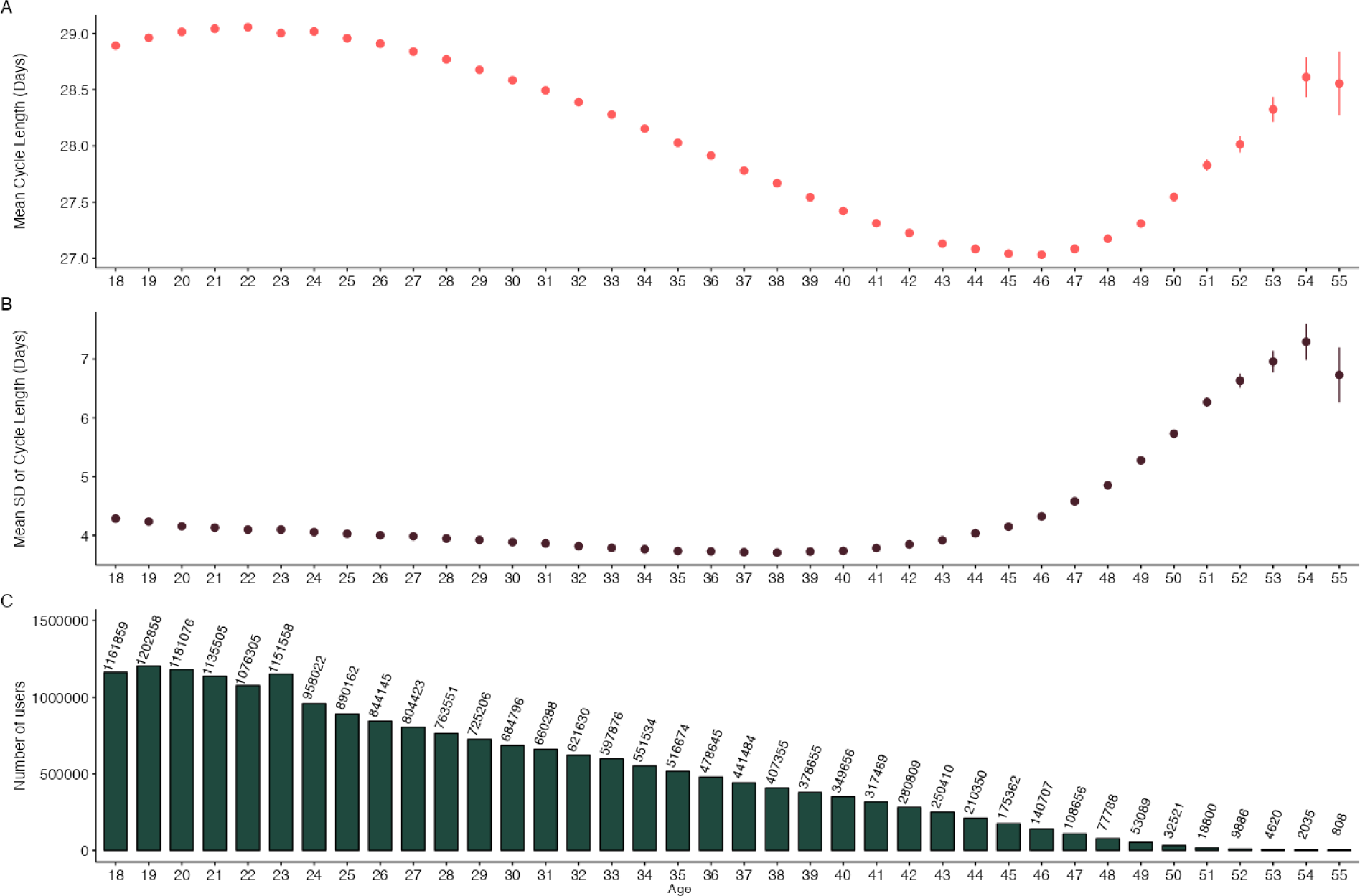
A) Mean cycle length and B) variation in cycle length as measured by standard deviation of cycle length and C) counts of number of users by age.

Cycle variability, as measured by standard deviation of cycle length for each user across the 12-month study period, also shows age associated differences (Table 1). In 18-25 year olds, the mean standard deviation of cycle length is 4.14 days, which is 1.1 times higher than the the mean standard deviation of cycle length in the 36-40 year olds (Mean SD = 3.72 days, Mean difference = 0.42 days, t=114.63, Cohen’s d=0.09). Compared to the minimum variability in the 36-40 age group, the oldest age group shows higher variability, at 6.52 days (Mean difference=2.80, t=111.43, Cohen’s d= 0.51). This pattern of gradually lower cycle variability from 18 to 37 years, before higher variability from the 37 onward is also seen when mean cycle variability is plotted by age (Figure 1B). Figure 1C indicates sample sizes by year of age.

The proportion of users with irregular cycles was highest in the 51-55 age band (44.7%), followed by the 46-50 age band (28.3%). Irregular cycles were least common in the 36-40 age (13.8%) band (Table 1).

### Period Length

The mean duration of periods experienced also differed by age, with the longest periods in the youngest and oldest age groups. Periods are longest in the 18-25 age group, with a mean duration of 5.20 days. Mean period length then lowers with advancing age to a minimum in the 41-45 age group, at 5.06 days (Mean difference= 0.22, t= 249.17, Cohen’s d= 0.24). However, mean period length becomes longer in the 51-55 year olds, compared to the 41-45 year old participants (Mean difference= 0.08, t= 15.6, Cohen’s d= 0.07). This U-shaped relationship is also revealed when mean period length is plotted by year of age. Figure 2A shows that mean period length is highest in 18 year olds, but gradually lowers until age 44, whereafter period length increases again in users aged 51-55.

**Figure 2.**
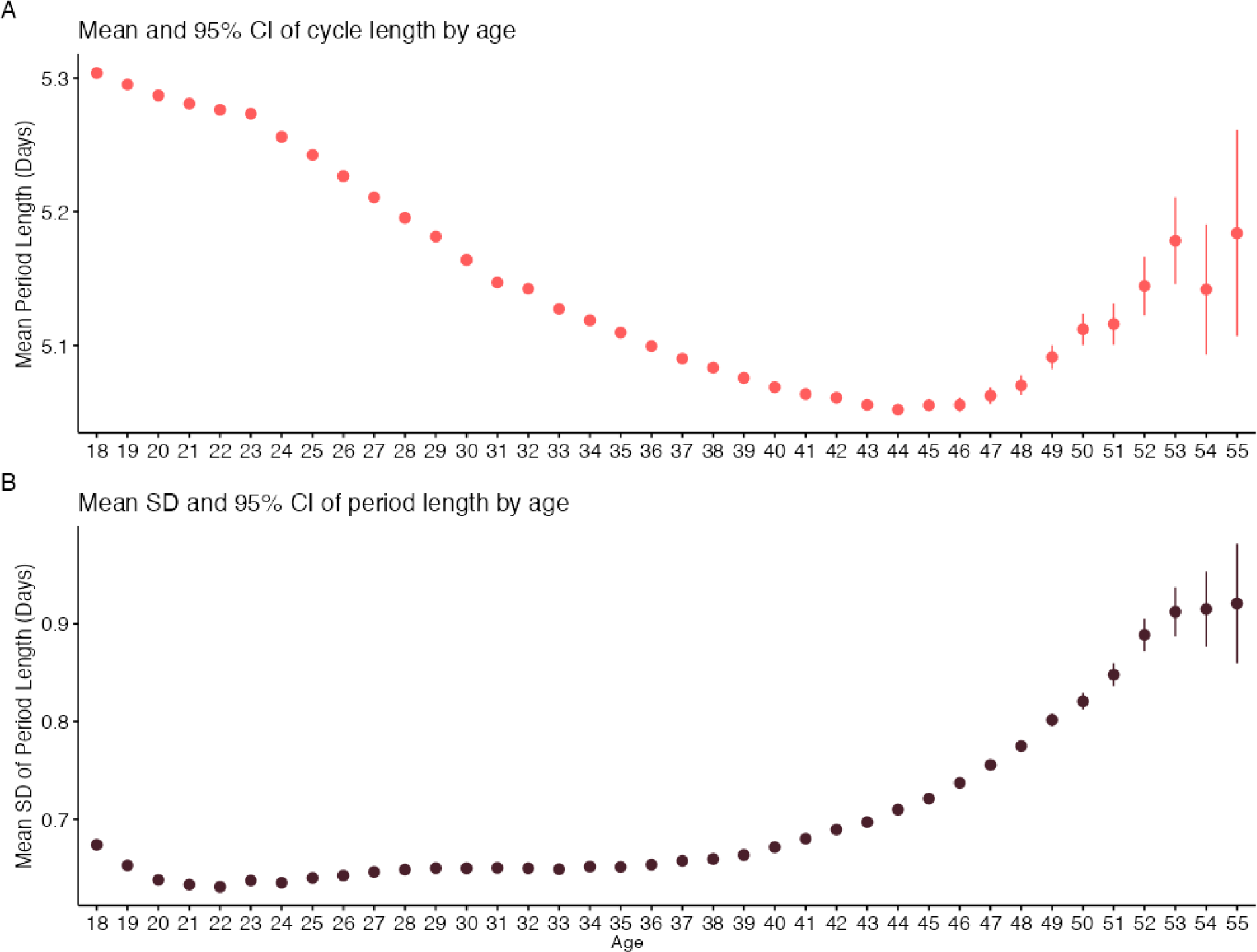
Plots of A) mean period length and B) variability in period length, as measured by standard deviation length, by age.

Variability in period length, as measured by the standard deviation of period length for each user across the 12-month study period, is lowest in users aged 26-30, and highest in the 51-55 age group. The mean difference between the 18-25 age group and 51-55 age group is 0.23 days (t= 67.81, Cohen’s d=0.31). This pattern is also seen when period variability is plotted by year of age. Figure 1B shows that variability in period length becomes smaller between 18 and 20 before becoming larger post age 20.

### Most frequent symptom logs by age

A total of 12,392,617 users had logged any of the symptoms of interest in the study time period. Out of the users that logged a symptom, 63,482/12,392617 (0.05%) were removed for an excessive number (more than five times the standard deviation of the mean of the total logs per user) of symptoms; a total of 12,329,135 users were included in the symptom level analysis.

The frequencies of symptom logs are presented in Supplementary Table 1. The top 10 symptoms logged by each age group are presented in Figure 3. For the 18-25, 26-30, 31-35, 36-40 and 41-45 age groups, the top three symptoms logged were cramps, breast tenderness and fatigue, though in the 26-30, 31-35, and 36-40 age groups fatigue was slightly more common than breast tenderness. In the 46-50 and 51-55 age groups, cramps, breast tenderness and headache were the top three most common symptoms logged.

**Figure 3.**
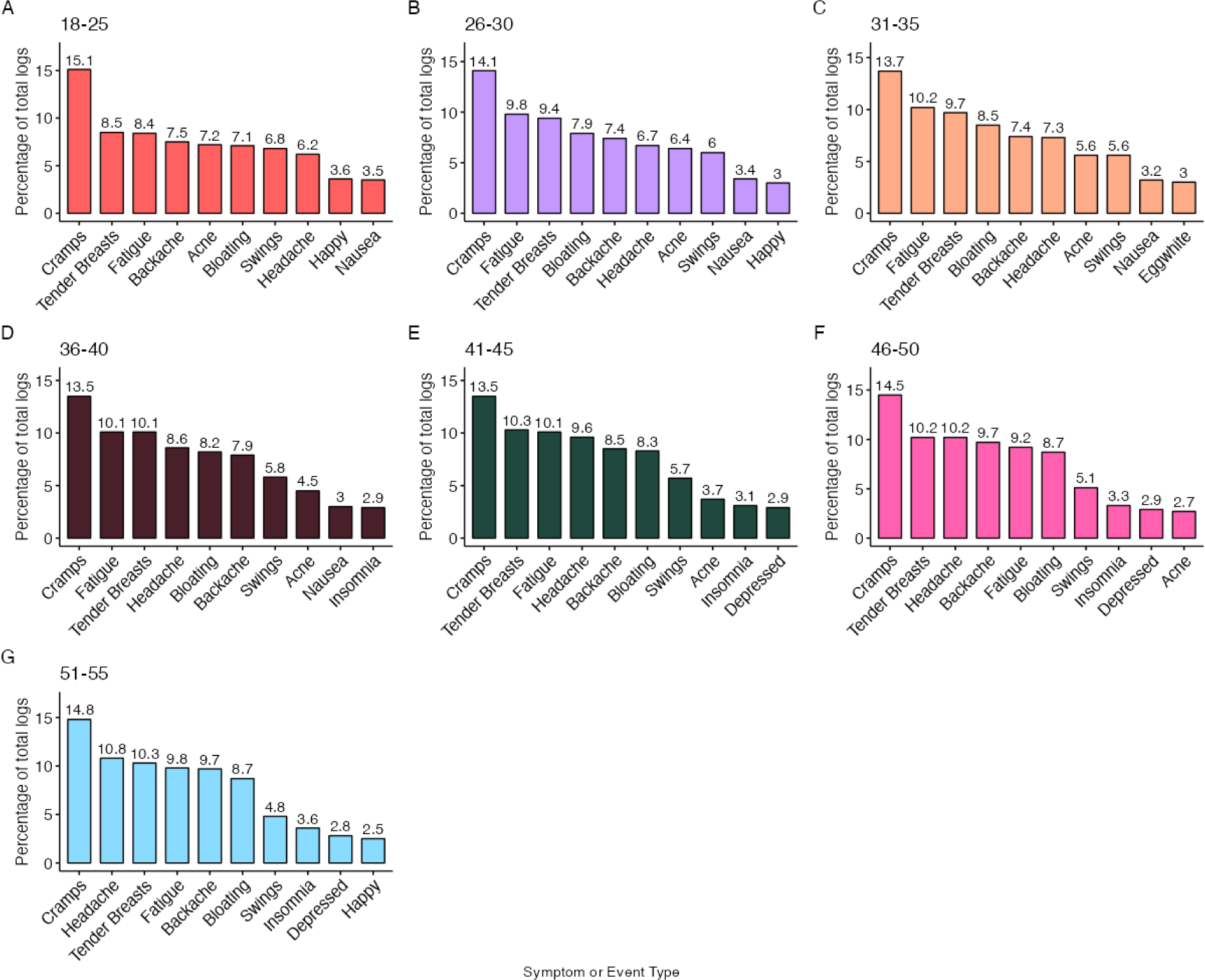
Percentage of total symptom logs by age band.

### Frequency of Symptoms

We assessed the mean number of logs by age for six somatic symptoms (Cramps, Tender Breasts, Headache, Fatigue, Backache and Acne) (Figure 4A). While logs of cramps and acne tend to decrease in frequency with advancing age, reports of headache, and backache tend to get more frequent in older participants. In contrast, reports of tender breasts and fatigue peak in the late twenties and early thirties.

**Figure 4.**
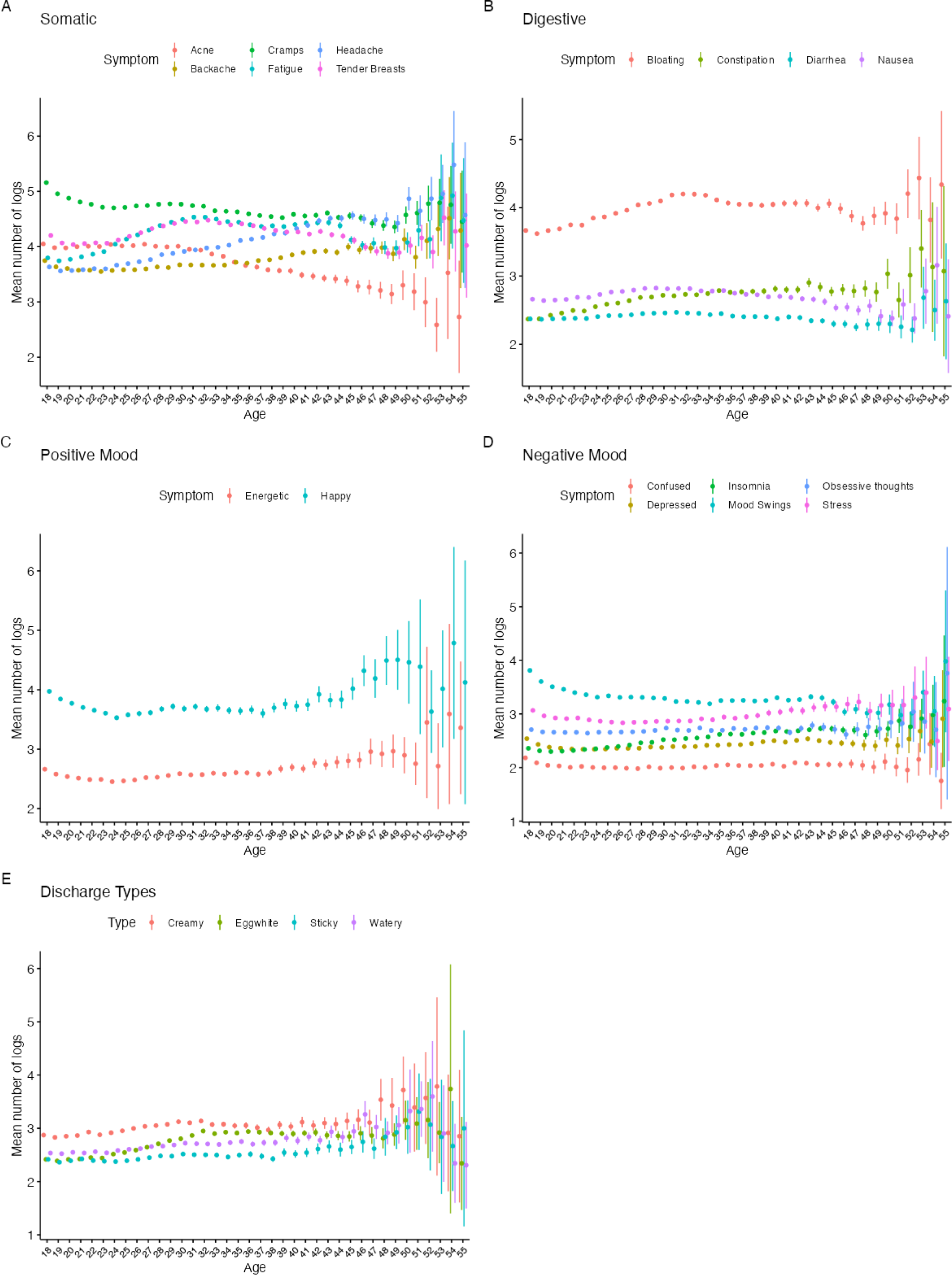
Mean number of symptom logs by age for A) Somatic, B) Digestive, C) Positive Mood, D) Negative Mood, and E) Vaginal Discharge.

For digestive symptoms (Figure 4B), including bloating, constipation and diarrhea, there is little difference in frequency of logging at different ages, but there is a larger number of logs of constipation at older ages, while overall, logs of bloating are 3.6 times higher than for constipation, 2.6 time higher than Diarrhea and 2.3 times higher than Nausea.

Mood symptoms also tend to remain fairly constant, albeit with a higher number of logs of happy or energetic mood at age 18 than in the 30-35 year olds. Logs of energetic mood are less frequent in older users, after age 49, and logs of happy mood peak around age 50 (Figure 4C). Overall negative mood logs also show little differences with age, though reports of mood swings tend to be highest in the youngest and oldest users, while stress and insomnia logs are higher in older than younger users (Figure 4D).

Finally, differences in vaginal fluid logs are seen between different ages. Logs of creamy, watery and sticky fluid remain constant until around age 47-48, where they become more frequent and variable. Frequency of egg white fluid logs show a peak from 32-47, before becoming variable after age 50 (Figure 4E).

## Discussion

In this paper we present the largest study to-date to investigate temporal characteristics of the menstrual cycle and self-reported symptom frequencies for a sample of over 19 million women aged 18-55. We found patterns of menstrual cycle and period length shortening and increasing variability with advancing age, and demonstrated age dependent patterns in self-reported menstrual cycle related symptoms.

The menstrual cycle does not remain static over the reproductive lifespan. Existing literature that has investigated the cycle in large epidemiological samples, or in mHealth samples, have clearly demonstrated that the cycle tends to get shorter, and more variable, with increasing age^32,33^. Broadly, these findings are replicated in this study, but the larger sample size, and wider age range allows for additional insights. Previous studies have demonstrated that cycle length increases between ages 18-23, before beginning a linear decrease in cycle length between the ages of 23 to 45 ^34^. However, here we can additionally demonstrate that after age 47 cycle length increases sharply until age 55, with a corresponding increase in cycle variability and cycle irregularity. Clinical guidelines are often structured around a median menstrual cycle length of 28 days, with timing of ovulation being approximately centered at around cycle day 14, and overall the mean cycle lengths presented here agree with these guidelines.

Similar to cycle length, the duration of bleeding days tends to decrease from a maximum in users aged 18 to a minimum around age 45, after which bleeding days length tends to increase. This pattern is mirrored by a general trend of increasing variability in bleeding days length with age. This increase in cycle and period length variability after age 45 is likely due to the menopausal transition, where cycles tend to become more variable. While the transition to menopause is generally associated with shorter cycles, there is evidence that cycle length actually increases near to menopause^5^, which is supported by our current findings, and is not shown in previous large mHealth studies investigating the menstrual cycle over narrower age bands^33,34^. Overall, our findings agree with these guidelines, mean cycle length is around 28 days in all age groups, but

Menstrual cycle related symptoms may differ in intensity or may even appear or disappear at different stages of the reproductive life span. Overall, the pattern of symptoms most frequently logged by users aged 51-55 is quite different to the pattern logged by users in the18-25 age band, with a greater number of logs for headache, fatigue, and backache for example in older users compared to younger users. These differences in the symptoms most frequently logged are also mirrored by distinct age related patterns in logging at an individual symptom level. A symptom like acne, which shows a general decreasing trend in logging with older age has a different pattern to headache, which shows a general increase in logging with age, and both display a different pattern compared to symptoms such as happy mood or energetic mood, which are most frequently logged in the youngest and oldest participants.

It is notable that around the age of 47 there are marked changes in cycle length and variability, along with changes in the frequency of symptom logs, with many symptoms showing a smaller or greater number of logs. For example, a decrease in bloating, nausea and negative mood logs such as stress, is mirrored by increases in energetic and happy mood logs and increased variability in vaginal discharge logs. While these differences in the frequency of logging of these symptoms around this age are clearly demonstrated, and are likely signs of the menopausal transition, they should be viewed in the context of a continuous change in the level of symptoms experienced over the lifespan. Traditionally, menopause has been viewed as a discrete period of change, but there are likely precursor changes that can be observed long before a woman knows they are perimenopausal.

Many menstruators feel that they do not have enough information about the menopause^14^, and a large proportion of individuals experience symptoms related to the menopause that are severe enough to lead them to seek out medical intervention^8–10^. Therefore, the results presented here are important in informing individuals about both the transition to menopause, and what to expect, and how the menstrual cycle changes over the lifespan. Understanding these differential experiences, and differences in what is normal at different life stages is important in a wide range of scenarios, from medical care, daily life, and family life. Maintaining health literacy through education, including through applications like Flo is important in ensuring that individuals know when they need medical attention, and for maintaining quality of life and satisfaction ^17,35,36^.

While this study, to our knowledge, is the largest study of its kind and the first to investigate age related menstrual cycle symptom patterns across such a wide age range, it does have limitations. Firstly, when reporting the start and end of a period, a user is initially offered to enter a duration of five days, but can change this duration. This may lead to a bias towards five day duration periods in the dataset. Secondly, it is known that ethnicity can affect the menstrual cycle. For example, individuals from Asia having an earlier onset of menopause on average than individuals from Europe ^37^. As part of Flo’s commitment to privacy, ethnicity information is not collected or stored by the app, meaning we are unable to investigate how ethnicity might change frequency of symptom logging, or cycle characteristics in this sample. While we excluded users who had set reminders to take contraception, we cannot be sure that all users included in the study are not taking hormonal contraception that could alter the cycle and symptoms. Similarly, we are not able to identify any users who are receiving hormonal therapy due to menopausal symptoms. Also, we also were not able to investigate frequency of logging of some symptoms that may be particularly related with perimenopause and menopause, such as hot flashes, night sweats or brain fog, since these symptoms are not available to log in the app. This, along with less frequent periods may mean that women who are experiencing the menopausal transition may be less inclined to engage with the app. This likely contributes to the age imbalance in our sample, where while we include over 1 million users older than 40 in our sample, the majority of participants are aged 18-25. Further research should endeavor to include classic menopause symptoms, along with extra information such as ethnicity and, if possible, lab based measurement of blood markers such as antimüllerian hormone, inhibin-B, and follicle stimulating hormone to obtain objective markers of ovarian reproductive aging. This would allow for better interpretation of how symptoms relate to ovarian aging, perimenopause and menopause.

## Conclusion

Here we present the results of the largest study investigating self reported menstrual cycle length, variability and associated symptoms in a sample of menstruating Flo *app* users aged 18-55. We demonstrate clear age-related trends in menstrual cycle and period length, in variability in these cycle indices as well as in symptom spectrum across the entire age range that spans from premenopause into the menopause transition. These patterns and differences in symptoms experienced are important in understanding the menstrual cycle at different life stages, and should be incorporated into educational and medical resources to better inform women about the menstrual cycle.

## Acknowledgements

The authors thank the participants who took part in the study. We would also like to thank Prof. Amanda Kowalski for reviewing the manuscript and providing advice and expertise.

## Author Contributions

The authors confirm contribution to the paper as follows: study conception and design: AC, LZ, LP; analysis and interpretation of results: AC, LZ, LP, AW, FG, AK CP; draft manuscript preparation: AC, CP, LZ, LP.

## Conflict of interest

AC, AW, LZ, and AK are employees of Flo Health. AC, LZ, AW, AK hold equity interests in Flo Health. LP, CP and FG are consultants for Flo Health.

## Ethical standards

The authors assert that all procedures contributing to this work comply with the ethical standards of the relevant national and institutional committees on human experimentation and with the Helsinki Declaration of 1975, as revised in 2008. The study protocol was approved by an independent ethics review board (Western Copernicus Group Independent Review Board - WCG IRB, number 20226050).

## Data Availability Statement

The data that support the findings of this study cannot be shared due to privacy restrictions.

## Notes

### Funding Statement

This study was funded by Flo Health

### Author Declarations

The study protocol was approved by an independent ethics review board (Western Copernicus Group Independent Review Board - WCG IRB, number 20226050).

