## Supplementary Material for "Chronicling menstrual cycle patterns across the reproductive lifespan with real world data"

*Supplementary Table 1. Frequencies of symptom logs.*

| Symptom | Frequency |
|---|---|
| Acne | 18643817 |
| Backache | 22203116 |
| Bloating | 22503741 |
| Confused | 4247955 |
| Constipation | 6239825 |
| Creamy | 7660203 |
| Depressed | 8346419 |
| Diarrhea | 8654778 |
| Cramps | 42467444 |
| Eggwhite | 7030332 |
| Energetic | 4987596 |
| Fatigue | 26951835 |
| Happy | 9663564 |
| Headache | 20305584 |
| Insomnia | 7040309 |
| Nausea | 9805926 |
| Obsessive Thoughts | 5695331 |

| | |
|---|---|
| Sticky | 4219391 |
| Stress | 7546165 |
| Mood Swings | 18484874 |
| TenderBreasts | 26663879 |
| Watery | 4289544 |
